# Clinical and genetic associations of asymmetric apical and septal left ventricular hypertrophy

**DOI:** 10.1101/2023.10.03.23296510

**Authors:** Victoria Yuan, Milos Vukadinovic, Alan C. Kwan, Florian Rader, Debiao Li, David Ouyang

## Abstract

Increased left ventricular mass has been associated with adverse cardiovascular outcomes including incident cardiomyopathy and atrial fibrillation. Such associations have been studied in relation to total left ventricular hypertrophy, while the regional distribution of myocardial hypertrophy is extremely variable and the clinical significant and genetic associations of such variability requires further study. Here, we use deep learning derived phenotypes of disproportionate patterns of hypertrophy, such as apical hypertrophy and septal hypertrophy, to study genome-wide and clinical associations in addition to and independent from total left ventricular mass within 35,268 UK Biobank participants. Adjusting for total left ventricular mass, apical hypertrophy is associated with elevated risk for cardiomyopathy and atrial fibrillation, and the risk for cardiomyopathy was increased for subjects with increased apical or septal mass even in the absence of global hypertrophy. We identified seventeen genome-wide associations for left ventricular mass, three unique associations with increased apical mass, and three additional unique associations with increased septal mass. Further studies are needed in multi-ethnic cohorts.

## Introduction

Left ventricular (LV) mass is associated with genetic variants of hypertrophic and dilated cardiomyopathy, and cardiovascular outcomes such as stroke, arrhythmias, and sudden cardiac death^1,2^. Increased total LV mass can reflect disease progression, such as a result of longstanding hypertension or valvular disease, while disproportionate or early onset hypertrophy can provide additional insight into genetic etiologies of myocardial hypertrophy. While there are known clinical patterns of asymmetric hypertrophy^3,4^, including septal and apical hypertrophy patterns^5–7^ in hypertrophic cardiomyopathy (HCM)^1,5,6^, there are few studies detailing the genetic determinants of focal or disproportionate hypertrophy^7^. Studies of asymmetric cardiomyopathy have been limited by cohort size and incomplete characterization by echoacardiography, such that the genetics of patterns of increased mass are not thoroughly explored^5,7,8^.

Deep learning-enabled high throughput evaluation of cardiac imaging opens an avenue for large-scale studies of cardiac phenotypes^9,10,11^. The ability to precisely phenotype cardiac imaging^12^ and combining underlying genetic information allows for the interrogation of a wide range of phenotypes and clinically meaningful traits. The United Kingdom BioBank (UKBB) initiative, particularly with the imaging cohorts which include cardiac magnetic resonance (CMR) imaging have been leveraged to understand the genetics of cardiovascular form and function^9,13,14^. With high fidelity imaging with CMR, deep learning can precisely and reproducibly evaluate imaging structures^10,12^. Previous studies have already evaluated LV wall thickness and mass^9,13,14^, finding strong associations with *TTN* and *CDKN1A* among other variants. In this work, we sought to build upon this foundation to evaluate the impact of focal hypertrophy.

Distinguishing between subtypes of LV hypertrophy may improve risk stratification^2^ and motivate targeted therapies^15^. Our study developed a deep learning algorithm to estimate total LV mass (LVM), apical LV mass, and septal mass from CMR images of 35,268 UKBB participants. These quantifications were used to examine the genetic basis for asymmetric LV mass and the associations and additional risk of isolated septal and apical hypertrophy with cardiovascular outcomes. Expression quantitative trait loci (eQTLs), gene set analysis, and chromatin interaction mapping were performed to further characterize the functional impact of these loci.

## Results

### Characterizing apical and septal mass in the UKBB cohort

We established a cohort of 35,268 individuals with measurable total LV, apex, and septum mass from the short axis CMR images (**Figure 1**). Subjects were excluded if images were low-quality or did not fully demonstrating the apex or septum (**Supplementary Figure 1**). LV septal and apical mass were both normally distributed with higher mean values observed among males (**Supplementary Figure 2)**. There was modest correlation between apical mass and total LV mass (r^2^ = 0.44) but a higher correlation between septal mass and LV mass (r^2^ = 0.74). Neither septal mass nor apical mass were significantly correlated with other traditional imaging measurements of the LV (**Figure 2**).

**Figure 1.**
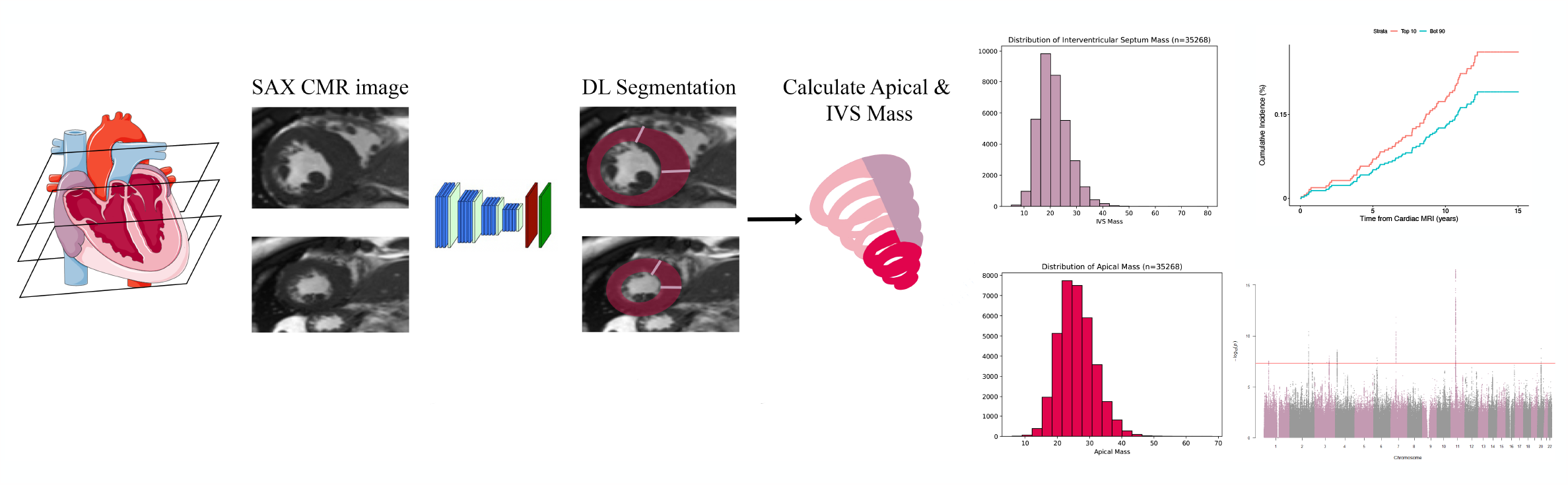
We characterized a cohort of 35, 268 participants for total LV, apical, and septal mass using deep learning. From these derived traits, we performed GWAS to identify genetic drivers of these phenotypes and analyzed the relationship between apical mass, septal mass, and incident cardiovascular disease.

**Figure 2.**
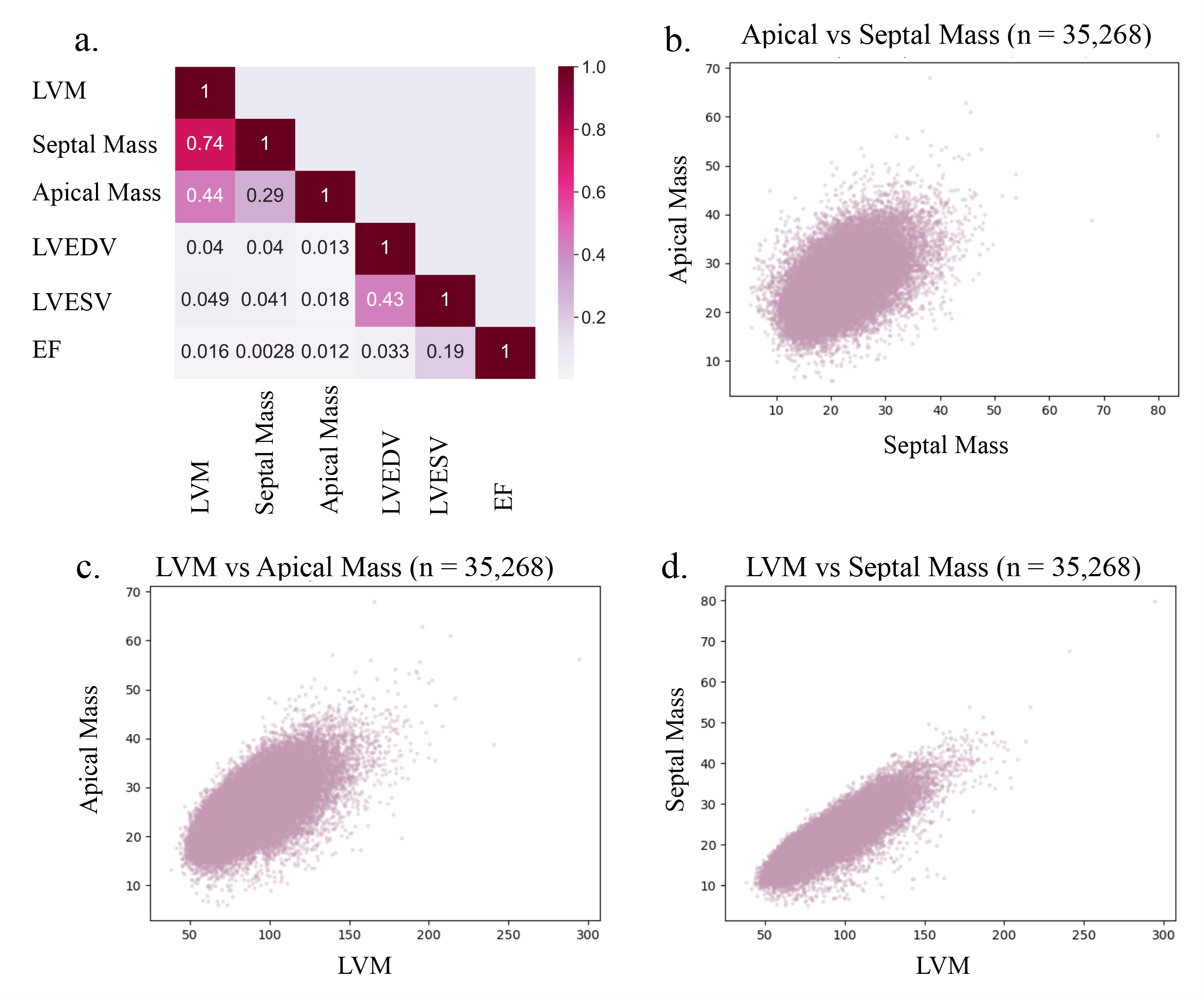
a) Heatmap showing the correlation between apical mass, septal mass, LVM, LVESV, LVEDV, and LVEF. Relationship between b) apical and septal mass, c) apical and LVM, and d) septal mass and LVM in the cohort.

### Focal hypertrophy’s association with incident cardiovascular disease

Given the correlation between focal regional mass and total left ventricular mass, we assessed the individual and combined contribution of increased mass and its association with incident cardiomyopathy (**Figure 3)**. Hypertrophy was defined as exceeding sex-specific 90^th^ percentile, and we identified subjects with global LVH, isolated apical hypertrophy without global hypertrophy, and isolated septal hypertrophy without global hypertrophy. Using a Cox proportional hazards model, subjects with global hypertrophy had the highest risk of cardiomyopathy (hazard ratio (HR) = 9.28, 95% confidence interval = [5.34, 16.11]). In parallel, isolated apical hypertrophy without the presence of global hypertrophy also conferred a higher risk of cardiomyopathy (HR = 2.69 [1.15, 6.28]) and isolated septal hypertrophy conferred a higher risk (HR = 4.41 [1.69, 11.53]). Our findings suggest that isolated regional hypertrophy confers excess risk for incident cardiomyopathy independent of global hypertrophy.

**Figure 3.**
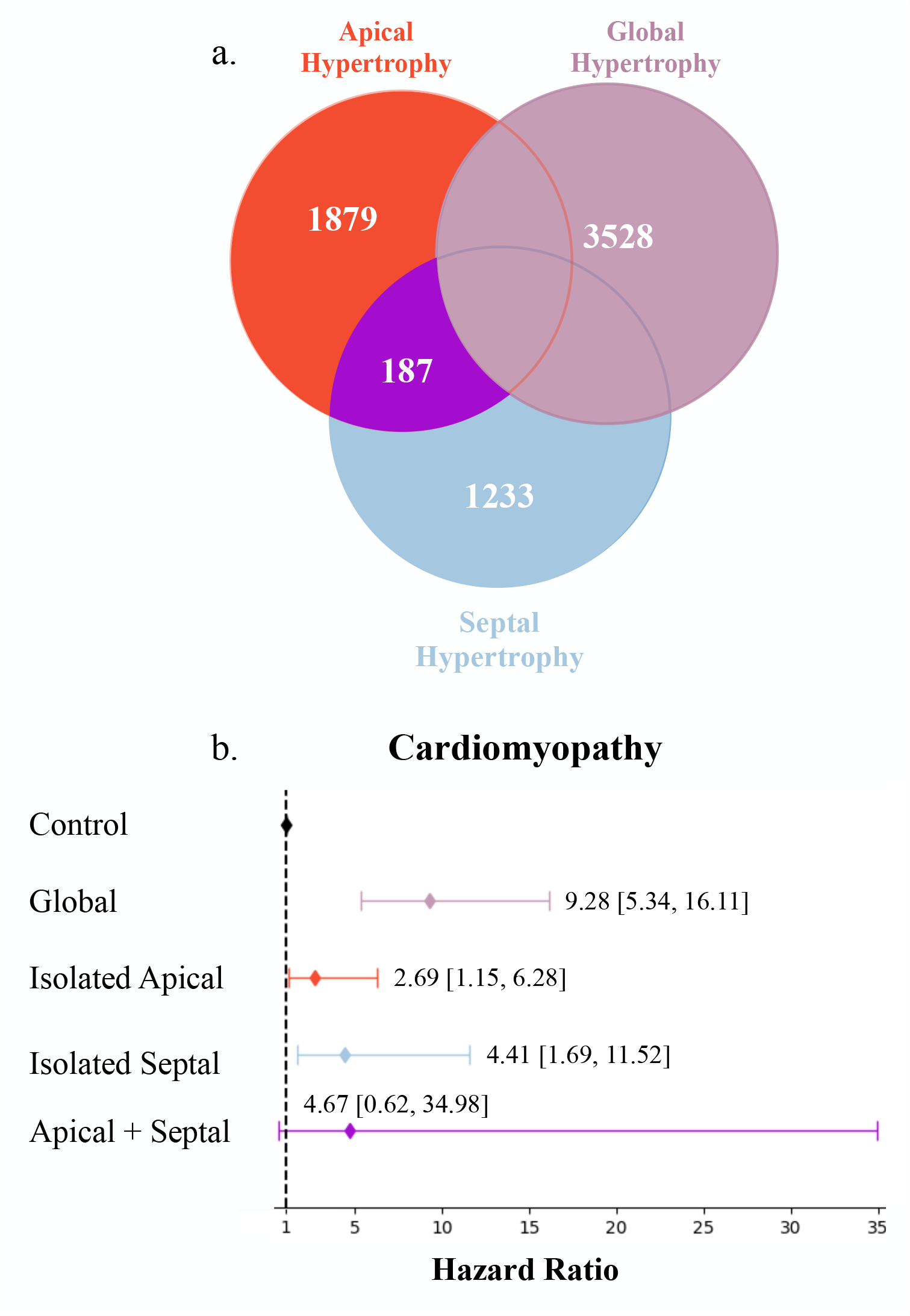
Each category of hypertrophy was defined as sex-specific 90^th^ percentile of mass. a) Venn diagram displaying global hypertrophy, isolated apical hypertrophy, isolated septal hypertrophy, and combined apical and septal hypertrophy (shaded in purple) b) Hazard ratios and 95% confident intervals for each phenotype with regards to cardiomyopathy. Subjects without hypertrophy (n = 28,441) served as the control group.

In addition to binary classification as hypertrophy, we investigated whether quantitative apical mass and septal mass are independent risk factors for incident cardiovascular disease. Using a Cox proportional hazards model, we found that increased apical, septal, and global mass predicts incident cardiomyopathy, atrial fibrillation, and myocardial infarction (Supplementary Figure 3). One standard deviation increase in LVM (22.29 g) was associated with a HR of 2.27 [2.02-2.55] increased risk for cardiomyopathy, and one standard deviation increase in apical mass (5.58 g) was associated with a HR of 2.89 [2.47, 3.51] for increased risk for cardiomyopathy. To test whether increased focal mass have independent predictive value beyond global LVM, we performed Cox analysis with models including both LVM and apical mass (M^LVM+apical^) and both LVM and septal mass (M^LVM+septal^). In M^LVM+apical^, in addition to a significant HR for global LVM, apical mass was also independent predictor of cardiomyopathy and atrial fibrillation. In contrast, in M^LVM+septal^, the HR for septal mass completely attenuated with adjustment for global LVM. Our results suggest septal mass predicts incident disease by proxying global LVM, while apical mass provides additive independent predictive value for incident cardiovascular disease.

### Genome-wide studies of CMR-derived global LVM, apical, and septal mass

We performed genome-wide association studies (GWAS) of apical and septal mass in 34,421 individuals who met CMR image and genetic quality criteria and compared the results with the genetic associations of global LVM (**Figure 4)**. For global LVM, seventeen independent variants reached genome-wide significance (**Table 1**), including many genes previously recognized to be associated with cardiovascular disease. *BAG3, BTN3A2* and other identified variants include those previously associated with hypertrophic cardiomyopathy, restrictive cardiomyopathy, mitral valve disease, and left ventricular dilation^16–25^ . GWAS of septal mass identified three unique genes (*HIVEP3, ESYT3*, and *HMGA1*) and nine genes shared with LVM. Among loci shared between the septal and LVM GWASs, *TTN* is an established gene for familial DCM, *GDF5* promotes cardiomyocyte survival, and loss of *GDF5* is associated with LV dilation and contractile dysfunction^24^. Of the genes unique to septal mass, *HIVEP3* is a transcription factor that is differentially methylated in HCM^26^, *ESYT3* is part of a genetic module that is differentially expressed in arrhythmogenic cardiomyopathy^27^, and *HMGA1* regulates cardiomyocyte growth with roles in concentric cardiac hypertrophy, myocardial infarction, and inflammation^28–30^.

**Table 1.**
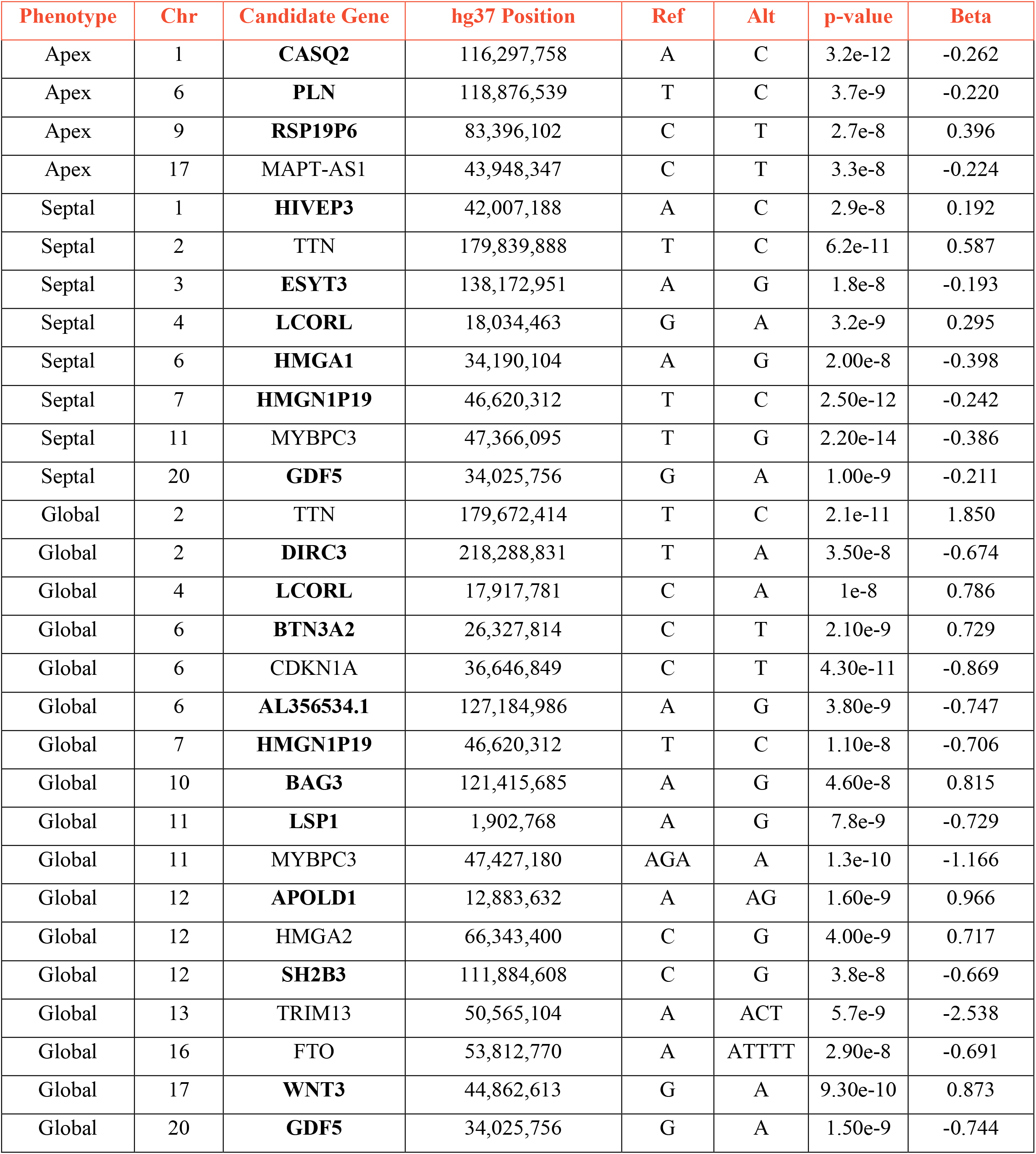
Genome-wide significant variants and candidate genes across all phenotypes, with novel genes in bold.

| Phenotype | Chr | Candidate Gene | hg37 Position | Ref | Alt | p-value | Beta |
| --- | --- | --- | --- | --- | --- | --- | --- |
| Apex | 1 | **CASQ2** | 116,297,758 | A | C | 3.2e-12 | -0.262 |
| Apex | 6 | **PLN** | 118,876,539 | T | C | 3.7e-9 | -0.220 |
| Apex | 9 | **RSP19P6** | 83,396,102 | C | T | 2.7e-8 | 0.396 |
| Apex | 17 | MAPT-AS1 | 43,948,347 | C | T | 3.3e-8 | -0.224 |
| Septal | 1 | **HIVEP3** | 42,007,188 | A | C | 2.9e-8 | 0.192 |
| Septal | 2 | TTN | 179,839,888 | T | C | 6.2e-11 | 0.587 |
| Septal | 3 | **ESYT3** | 138,172,951 | A | G | 1.8e-8 | -0.193 |
| Septal | 4 | **LCORL** | 18,034,463 | G | A | 3.2e-9 | 0.295 |
| Septal | 6 | **HMGA1** | 34,190,104 | A | G | 2.00e-8 | -0.398 |
| Septal | 7 | **HMGN1P19** | 46,620,312 | T | C | 2.50e-12 | -0.242 |
| Septal | 11 | MYBPC3 | 47,366,095 | T | G | 2.20e-14 | -0.386 |
| Septal | 20 | **GDF5** | 34,025,756 | G | A | 1.00e-9 | -0.211 |
| Global | 2 | TTN | 179,672,414 | T | C | 2.1e-11 | 1.850 |
| Global | 2 | **DIRC3** | 218,288,831 | T | A | 3.50e-8 | -0.674 |
| Global | 4 | **LCORL** | 17,917,781 | C | A | 1e-8 | 0.786 |
| Global | 6 | **BTN3A2** | 26,327,814 | C | T | 2.10e-9 | 0.729 |
| Global | 6 | CDKN1A | 36,646,849 | C | T | 4.30e-11 | -0.869 |
| Global | 6 | **AL356534.1** | 127,184,986 | A | G | 3.80e-9 | -0.747 |
| Global | 7 | **HMGN1P19** | 46,620,312 | T | C | 1.10e-8 | -0.706 |
| Global | 10 | **BAG3** | 121,415,685 | A | G | 4.60e-8 | 0.815 |
| Global | 11 | **LSP1** | 1,902,768 | A | G | 7.8e-9 | -0.729 |
| Global | 11 | MYBPC3 | 47,427,180 | AGA | A | 1.3e-10 | -1.166 |
| Global | 12 | **APOLD1** | 12,883,632 | A | AG | 1.60e-9 | 0.966 |
| Global | 12 | HMGA2 | 66,343,400 | C | G | 4.00e-9 | 0.717 |
| Global | 12 | **SH2B3** | 111,884,608 | C | G | 3.8e-8 | -0.669 |
| Global | 13 | TRIM13 | 50,565,104 | A | ACT | 5.7e-9 | -2.538 |
| Global | 16 | FTO | 53,812,770 | A | ATTTT | 2.90e-8 | -0.691 |
| Global | 17 | **WNT3** | 44,862,613 | G | A | 9.30e-10 | 0.873 |
| Global | 20 | **GDF5** | 34,025,756 | G | A | 1.50e-9 | -0.744 |

**Figure 4.**
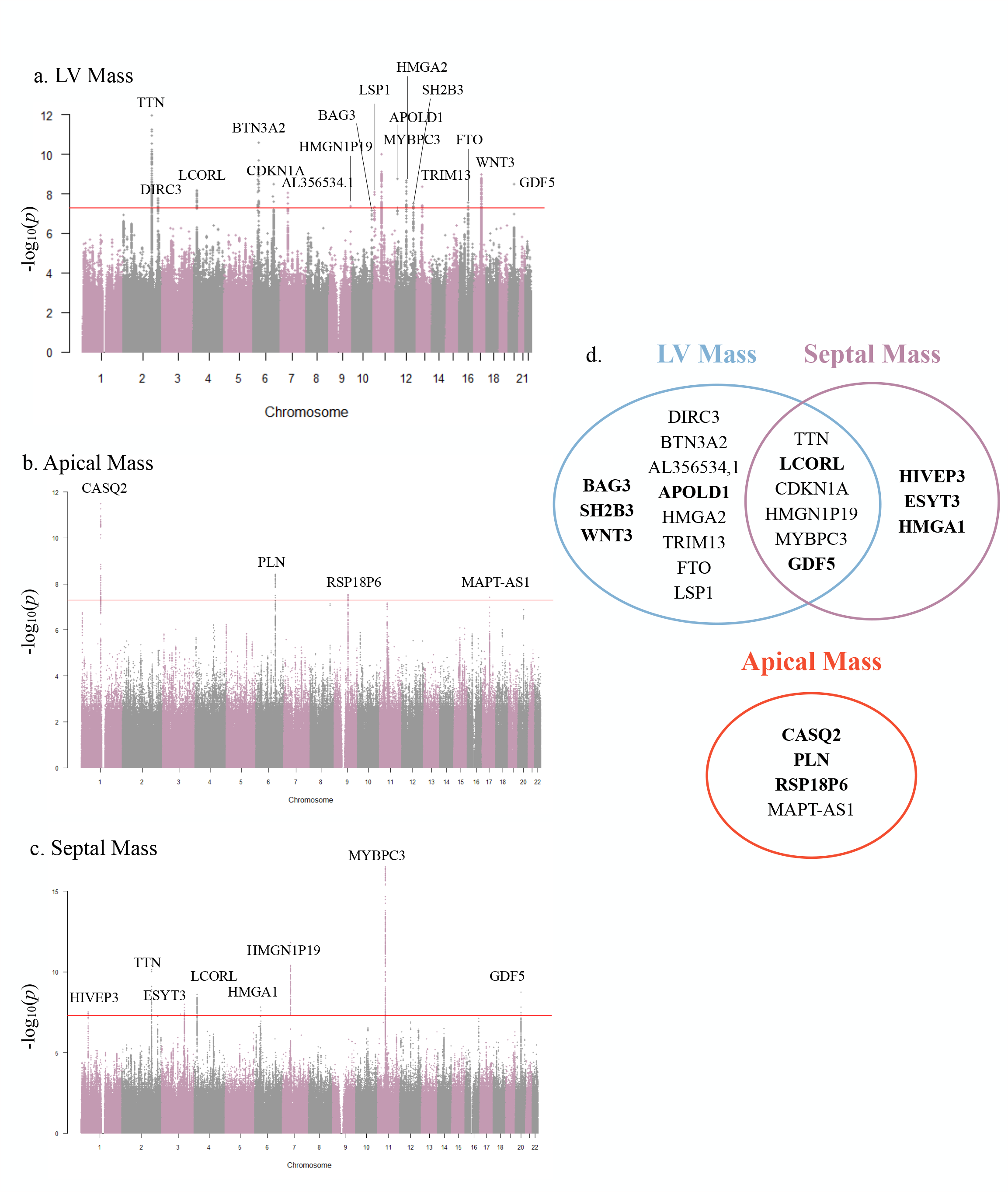
Manhattan plots for genome-wide association studies of a) LVM b) apical mass and c) septal mass. The horizontal red line represents a genome-wide significant p-value of 5e-8. d) Significant hits are compared across the three phenotypes, with novel loci in bold.

For apical mass, we identified three distinct loci that were not previously associated with total LVM or septal mass and that have not been previously associated with apical hypertrophy^3,31^. *CASQ2* encodes the protein calsequestrin, with pathologic variants results in aberrant calcium release from the sarcoplasmic reticulum, contractile dysfunction, dilated cardiomyopathy, and catecholaminergic ventricular arrhythmia without structural heart disease^32,33^. *PLN* plays a causal role in dilated, hypertrophic, arrhythmogenic, and familial cardiomyopathies^34,35^. *MAPT-AS1*, which has been previously correlated with LVM^9^ and cross-sectional area of the septal wall^36^, regulates sarcomere assembly and function^37^; its dysfunction has been attributed to heart failure with preserved ejection fraction^38^.

### Functional investigation of markers for apical and septal mass

We leveraged eQTL analysis and Hi-C chromatin mapping to annotate the functions of significant variants of apical and septal mass. eQTL analysis of apical mass variants reveals that the lead variant is associated with increased expression of *CASQ2* in the left ventricle, atrial appendage, and coronary artery (**Supplementary Table 1**). With Hi-C chromatin mapping, we also identified interactions between *CASQ2* and *NHLH2*, a gene with elevated expression in DCM^39^, and between *CASQ2* and *VANGL1*, which is involved in the development of cardiac outflow tracts^40^. Additionally, *PLN* interacts with *ASF1A*, a gene involved with cardiogenic mesoderm development and associated with ventricular and atrial septal defects. The lead variant for *PLN* is associated with increased expression in the aorta and atrial appendage. *MAPT-AS1* is associated with increased expression in the LV and has chromatin contacts with *KANSL1*, which modulates congenital heart defects, and *WNT3*, which regulates cardiomyocyte differentiation and cardiac mesoderm^21–23,41^. To further investigate the physiological function of genes associated with apical mass, we performed gene set analysis. We found strong associations with networks for cardiomyocyte contraction by calcium ion signaling (p = 4.01e-5), regulation of sequestered calcium ion release for cardiac muscle contraction (p = 5.25e-5), and cell communication by electrical coupling (p = 6.54e-6).

Applying this analysis to variants governing septal mass, we found that *HIVEP3* interacts with *EDN2*, a risk factor for atrial fibrillation in individuals with hypertrophic cardiomyopathy^42^. *GDF5*, which is shared between the total LV and septal mass phenotypes, has chromatin contacts with *MYH7B*, which in turn activates the CaMK signaling pathway involved in the pathophysiology of HCM^43^. *ESYT3* is associated with enriched expression in the coronary arteries (**Supplementary Table 1**). Finally, with gene set analysis, we discovered that genes governing septal mass are significantly associated with pathways for the morphogenesis and development of the atria (p_Bon_ = 0.0087), septum (p_Bon_ = 0.0122), and striated muscle (p_Bon_ = 0.0042).

## Discussion

In this study, we leveraged deep learning to identify novel genetic variants of CMR-derived total LV, apical, and septal mass in over 35,000 individuals and established their relationship to incident cardiovascular disease. Notably, apical and septal hypertrophy without increased total LVM both conferred independent excess risk for cardiomyopathy, and apical mass had unique genetic loci compared to prior associations with septal and total LVM. Our findings emphasize the diagnostic importance of focal hypertrophy, which may confer independent as well as additive risk for incident cardiovascular disease. Through downstream *in silico* analysis, we found that genes for apical and septal mass govern cardiovascular structure, function, and implicated in cardiomyopathy, further providing evidence of unique genetic and risk profiles for patterns of hypertrophy.

Whether evaluated as categorically as hypertrophy or based on quantitative mass, both increased apical and septal mass resulted in an increased risk for incident cardiomyopathy that was only partially attenuated by global LV hypertrophy. Subjects with isolated apical cardiomyopathy and isolated septal hypertrophy have a higher risk of cardiomyopathy even without global hypertrophy as defined by current clinical protocols^44^. The attenuation of the effect of septal mass on incident cardiovascular disease by global LVM parallels the overlapping loci in our genetic association studies for septal mass and global LVM. Nevertheless, we identify three genes unique to septal mass, emphasizing that septal hypertrophy may be governed by novel genes in addition to those classically attributed to global hypertrophy, such as *TTN*. Additionally, we uncover variants unique to apical mass, which have not been previously associated with apical hypertrophic cardiomyopathy^3,31^. Notably, *CASQ2* and *PLN* maintain cardiac calcium homeostasis, and their dysfunction increases the risk of atrial fibrillation^45,46^, which is a common complication in apical HCM^47^. These unique loci suggest that apical hypertrophy may have a genetically distinct pathophysiology, which may contribute to its different clinical outcomes and treatment from other types of HCM. Our findings highlight the significance of focal hypertrophy to cardiovascular pathophysiology^5–8,48^ and reaffirm the value of characterizing distinct subtypes of LV hypertrophy.

While smaller clinical cohorts are often underpowered to assess a difference in clinical outcomes, the presence of focal hypertrophy in the UKBB imaging cohort highlights the ability to identify unique subpopulations and their subsequent risk for incident cardiovascular disease. Understanding these patterns of regional hypertrophy may paint a fuller picture of cardiovascular disease outcomes and risk. Limitations of our study include the lack of replication across other cohorts. Although the UKBB cohort is of mixed-ancestry, European descent predominates, and our results may not be generalizable for individuals of other ancestries.

In conclusion, we analyzed the genotypic and clinical significance of apical, septal, and LV mass in over 35,000 UKBB participants. Our results identify twelve loci of clinically observed increased left ventricular mass, with three unique loci to increased apical mass. Our findings suggest that increased apical mass is a significant additional independent risk factor for cardiomyopathy and atrial fibrillation. Such results might be relevant to existing work in characterizing subtypes of hypertrophic cardiomyopathy, as we find distinct risk profiles for incident cardiovascular disease based on pattern of focal hypertrophy.

## Methods

### UK Biobank

The UK Biobank (UKBB) is a prospective study with 502,461 participants from 40 to 60 years of age. Detailed non-imaging data, such as genotyping, diagnoses, and environmental factors are also included. Our analysis focused on the 45,361 individuals who underwent cardiac magnetic resonance (CMR) imaging.

### Measurement of apical and interventricular septal mass

We leveraged a fully convolutional neural network to segment the end-diastolic short-axis CMR images of 45,361 individuals in the UK Biobank (UKBB). Participants were excluded based on image quality and visualization of the ventricles and apex (**Supplemental Figure 1**). We established our final study cohort (n=35,268) to measure apical and interventricular mass. Apical mass was calculated by isolating the pixels corresponding to the LV myocardial wall in the last four slices. The pixels were then summed to obtain apical area, then multiplied by slice thickness and slice gap to calculate apical volume. Finally, the volume was multiplied by myocardial density (1.055 g/cm^3^) to obtain apical mass. For each slice, we defined the septum as the segment of the LV myocardium bounded by the insertion points of the right ventricle wall. The pixels were summed to calculate the septal area, then multiplied by slice thickness, slice gap, and myocardial density to measure septal mass.

### Testing for associations with incident disease

We analyzed associations between CMR-derived LVM, apical mass, and septal mass with cardiomyopathy, atrial fibrillation, and acute myocardial infarction. We also examined the relationship between these outcomes and global, apical, and septal hypertrophy. Based on current clinical definitions, we defined global hypertrophy as individuals with overall LVM above the sex-specific 90^th^ percentile^49^. Similarly, apical hypertrophy was defined as individuals with 90^th^ percentile of apical mass and septal hypertrophy as the 90^th^ percentile of septal mass. Cox proportional hazards analysis was performed adjusted for sex, age pulse rate, and hypertension. Incident disease was identified using the reported International Classification of Disease 9^th^ and 10^th^ codes available in UKB starting from the time of CMR acquisition until the date of the first incident, death, or the last follow-up.

To analyze apical, septal and LV mass as continuous variables, we leveraged Cox proportional hazards testing in 3 different models. The first model investigated each mass phenotype individually as independent risk factors while adjustments for sex, age at MRI acquisition, pulse rate, and hypertension. The second model jointly analyzed apical mass and LVM, with the same adjustments as the first model. The third model jointly analyzed septal mass and LVM. To test associations between cardiomyopathy and ventricular hypertrophy, we created seven different cohorts.

### Genome-wide Association Study

Given our cohort of 35,268 individuals, 847 were removed due to incomplete or low-quality genetic data. We then performed GWAS of total LVM, apical mass, and septal mass on 34,421 individuals with BOLT-LMM v2.3.4^50^, which uses a Bayesian mixture prior as a random effect to fit a linear mixed model. We utilized the UKB imputed genotype calls in BGEN v1.2 format. Variants were required to have a minor allele frequency (MAF) ≥ 0.01, and imputed variants had an INFO score ≥ 0.3. Our model was adjusted for age at CMR imaging, sex, and the first 10 principal components of genetic ancestry. Variants were considered statistically significant at the standard genome-wide significance level of p = 5e-8.

Independent significant SNPs were then defined as SNPs that met the threshold for genome-wide significance, had an r^2^ > 0.6, and were located at least ±500 kb away from each other.

### Functional annotation of significant loci

We utilized FUMA v1.6.0^51^ to investigate eQTL, 3D chromatin interaction mapping, and gene set analysis to better understand the function of genome-wide significant variants. First, using GTEx version 8 eQTL tissue data, we evaluated the relationship between SNPs of genome-wide significance with cardiac gene expression in the aorta, atrial appendage, and LV tissue. GTEx contains pre-calculated false discovery rates (FDR) for gene-tissue pairs. FUMA defines significant eQTLs as SNP-gene pairs with p < 0.05 and gene-tissue pairs with an FDER ≤ 0.05. Second, 3D chromatin interaction mapping was performed using pre-processed Fit-Hi-C datasets of the LV, right ventricle, and aorta. One end of a signification interaction is defined by the independent, significance SNP and those in linkage disequilibrium with it. The other end of the significant regions is the mapped gene reported in the Results section. Finally, we performed gene set analysis using 10,678 curated gene sets annotated with Gene Ontology terms; Bonferroni correction was performed for all gene sets. Significant gene sets were those with a p_Bon_ < 0.05.

## Data Availability

UK Biobank data is available with an approved study protocol.

https://www.ukbiobank.ac.uk/

## Supplementary Information

**Supplementary Figure 1.**
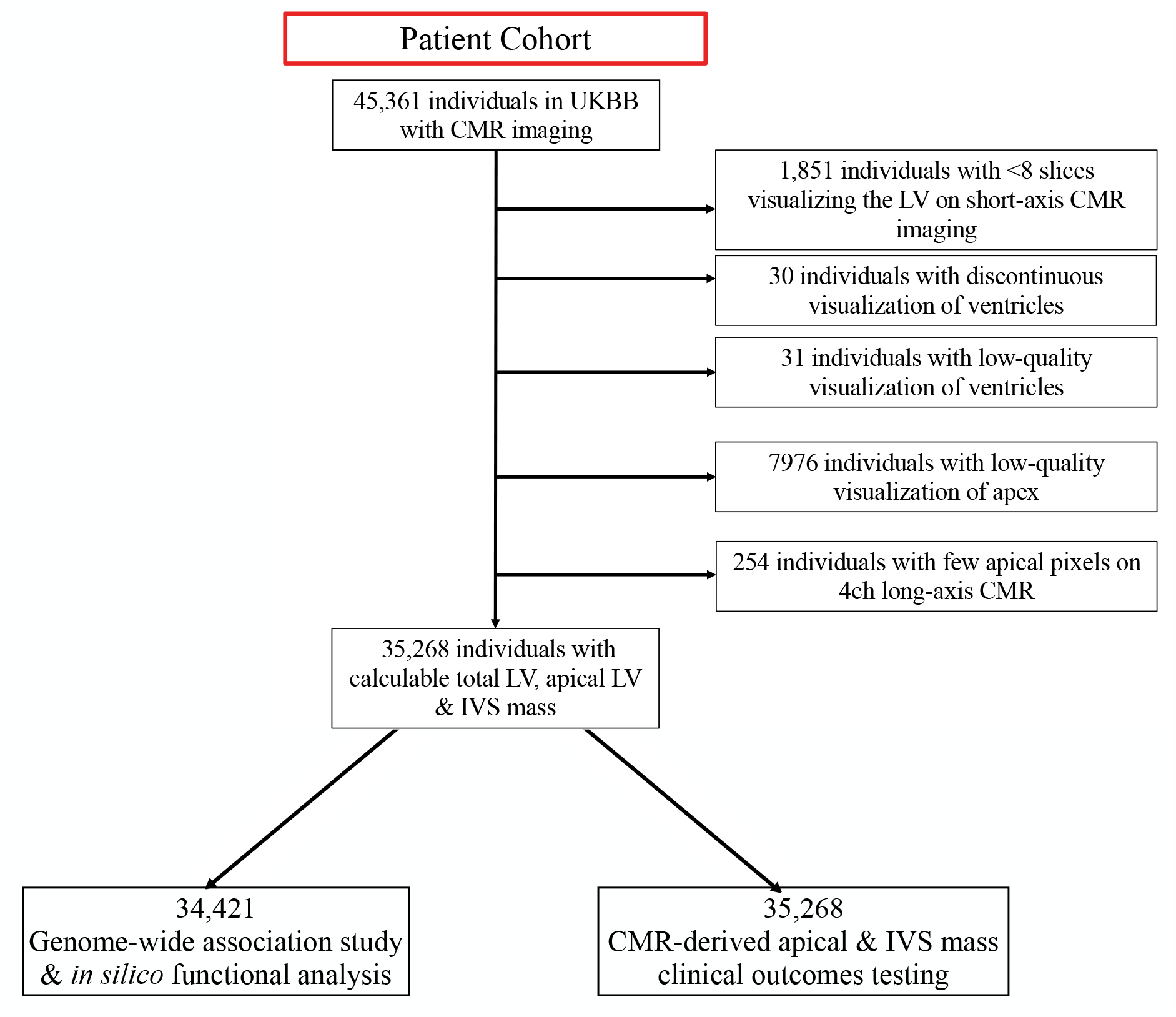
Flow diagram to create the cohort for genetic and phenotypic analysis

**Supplementary Figure 2.**
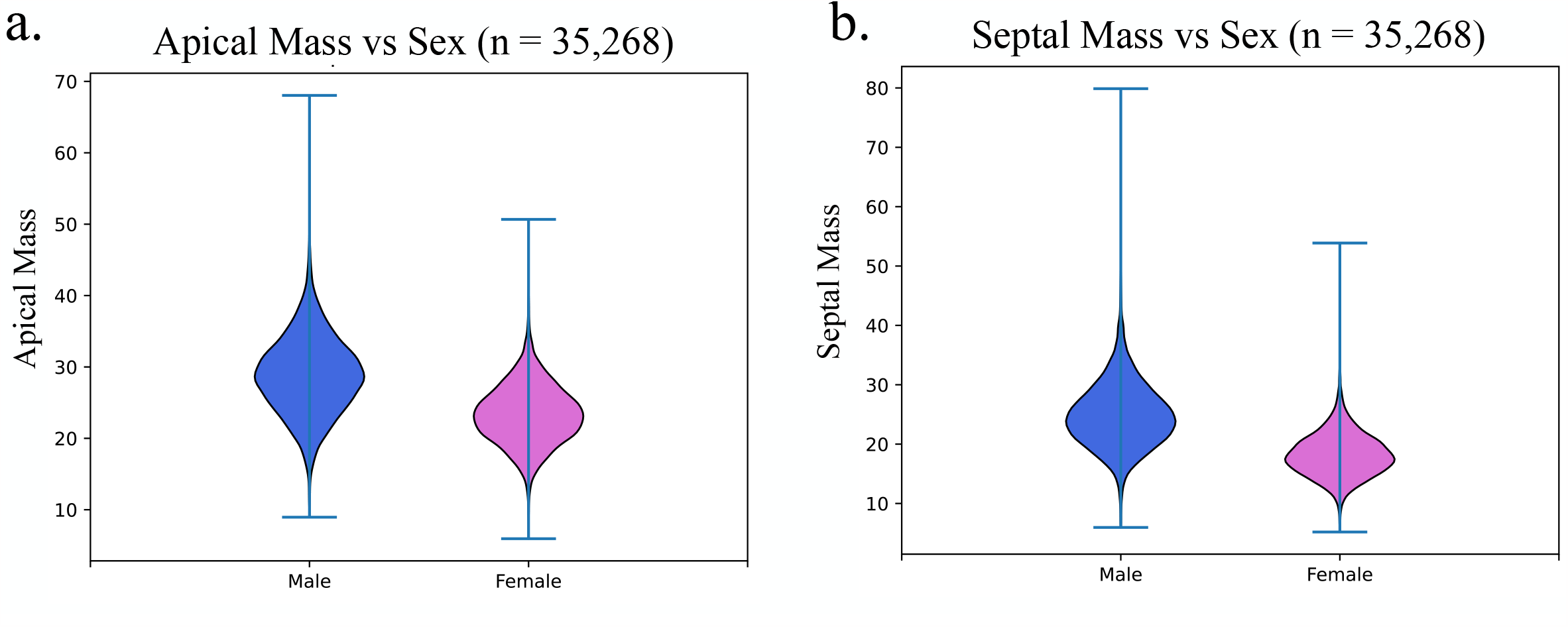
Distributions of a) apical mass and b) septal mass versus sex.

**Supplementary Figure 3.**
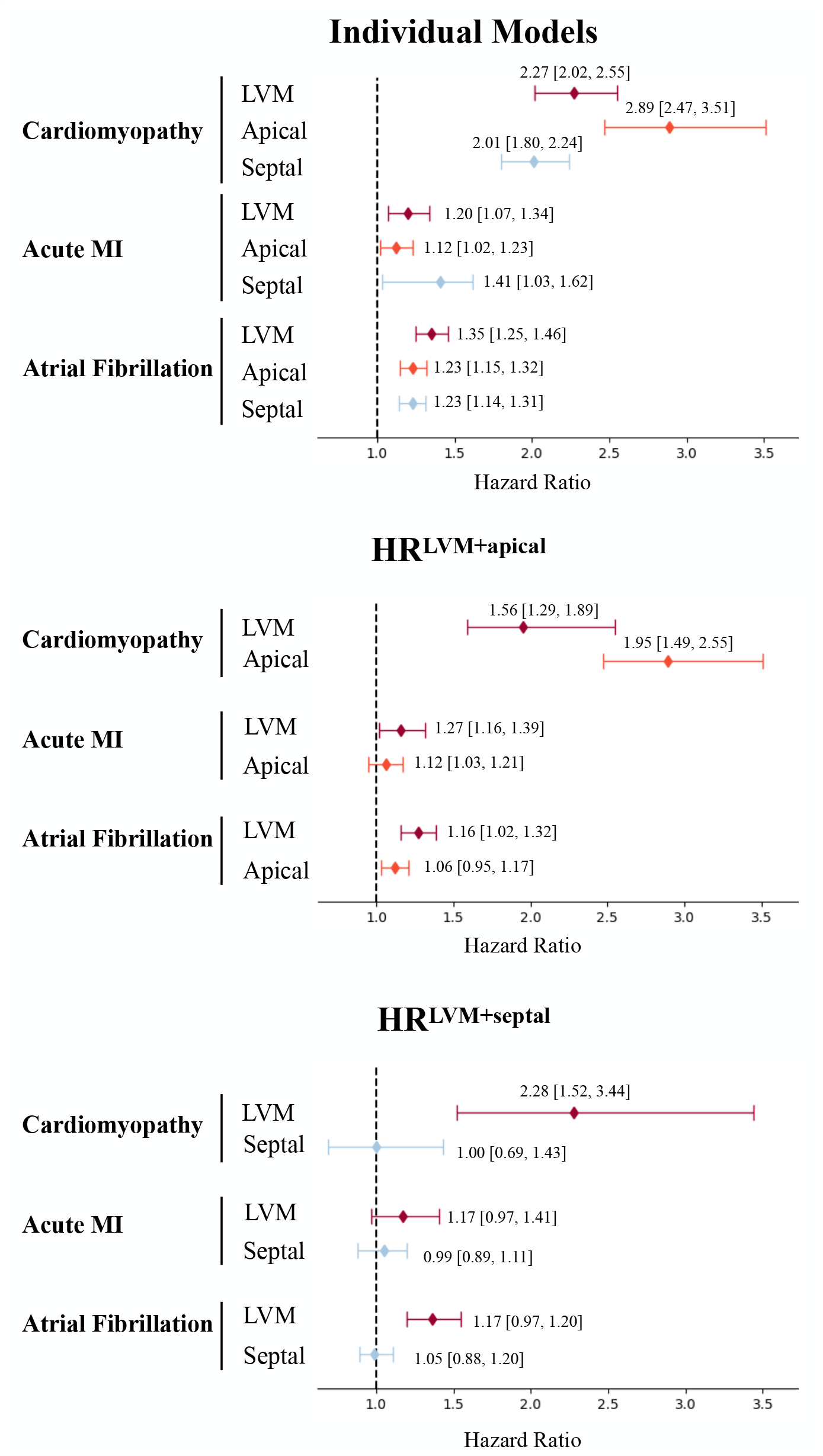
Hazard ratios with 95% confidence intervals for cardiomyopathy, atrial fibrillation, and acute MI. Individual models include the risk conferred by LVM, apical mass, and septal mass when investigated individually as independent variables in models adjusted for sex, age at MRI, BMI, heart rate, and hypertension. Model 2 includes apical mass, LVM, sex, age at MRI, BMI, heart rate, and hypertension, and Model 3 includes septal mass, LVM, sex, age at MRI, BMI, heart rate, and hypertension.

**Supplementary Table 1.**
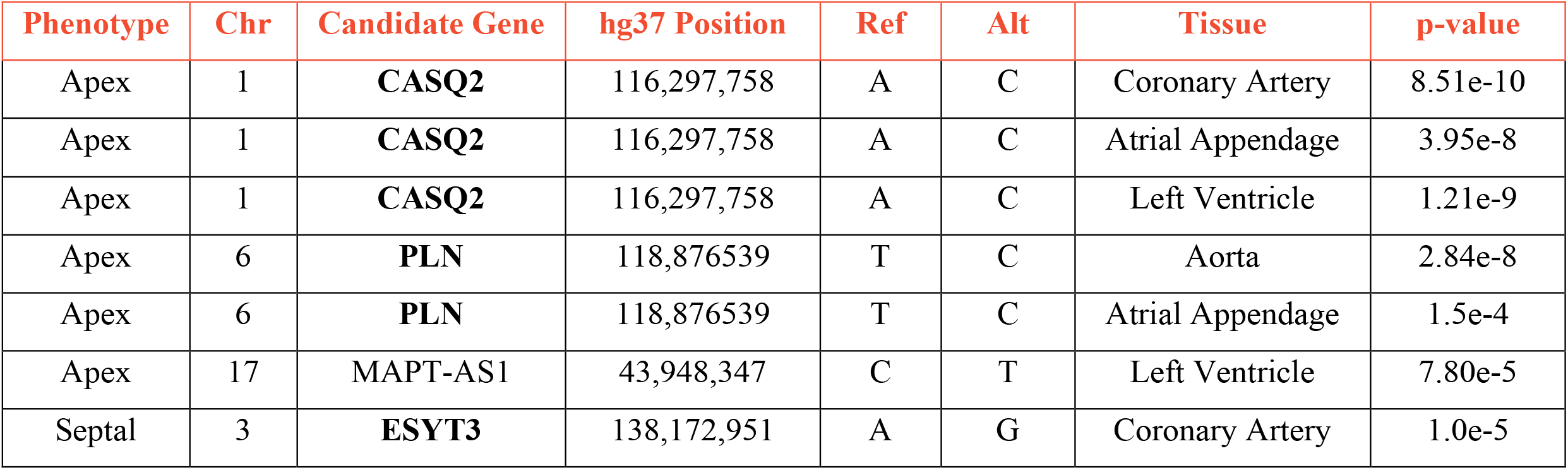
Results of eQTL analysis using FUMA for lead variants in the GWAS of apical and septal mass.

