## Supplementary figures and images for "Clinical and genetic associations of asymmetric apical and septal left ventricular hypertrophy"

### Supplemental Figure 2

**Supplementary Figure 2.** Distributions of a) apical mass and b) septal mass versus sex.

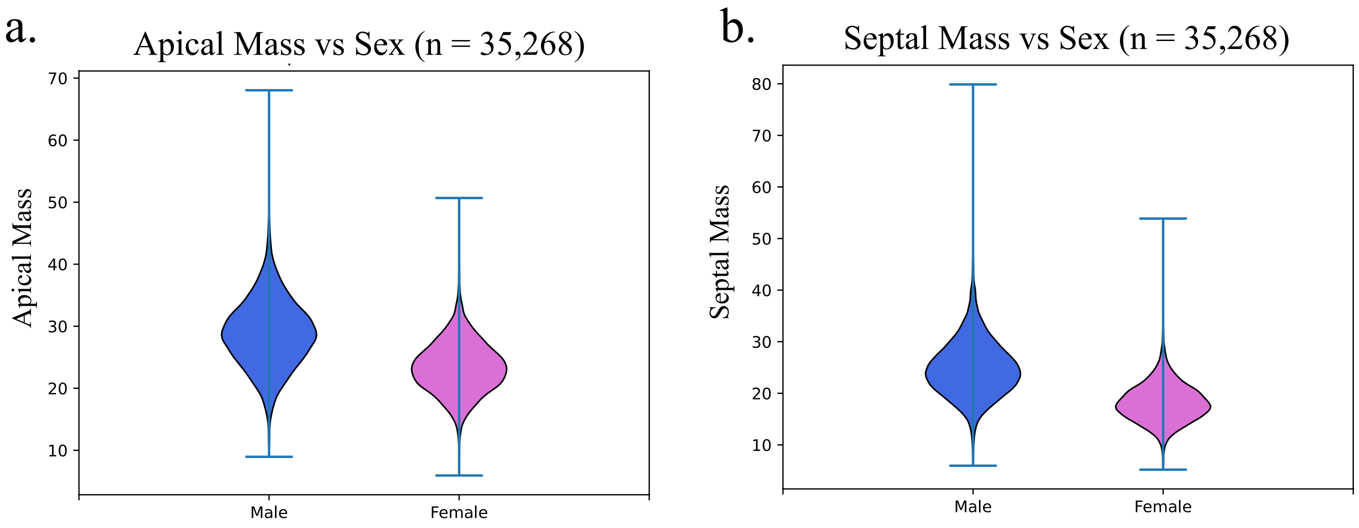
