## Supplemental Figure 3 for "Clinical and genetic associations of asymmetric apical and septal left ventricular hypertrophy"

**Supplementary Figure 3.** Hazard ratios with 95% confidence intervals for cardiomyopathy, atrial fibrillation, and acute MI. Individual models include the risk conferred by LVM, apical mass, and septal mass when investigated individually as independent variables in models adjusted for sex, age at MRI, BMI, heart rate, and hypertension. Model 2 includes apical mass, LVM, sex, age at MRI, BMI, heart rate, and hypertension, and Model 3 includes septal mass, LVM, sex, age at MRI, BMI, heart rate, and hypertension.

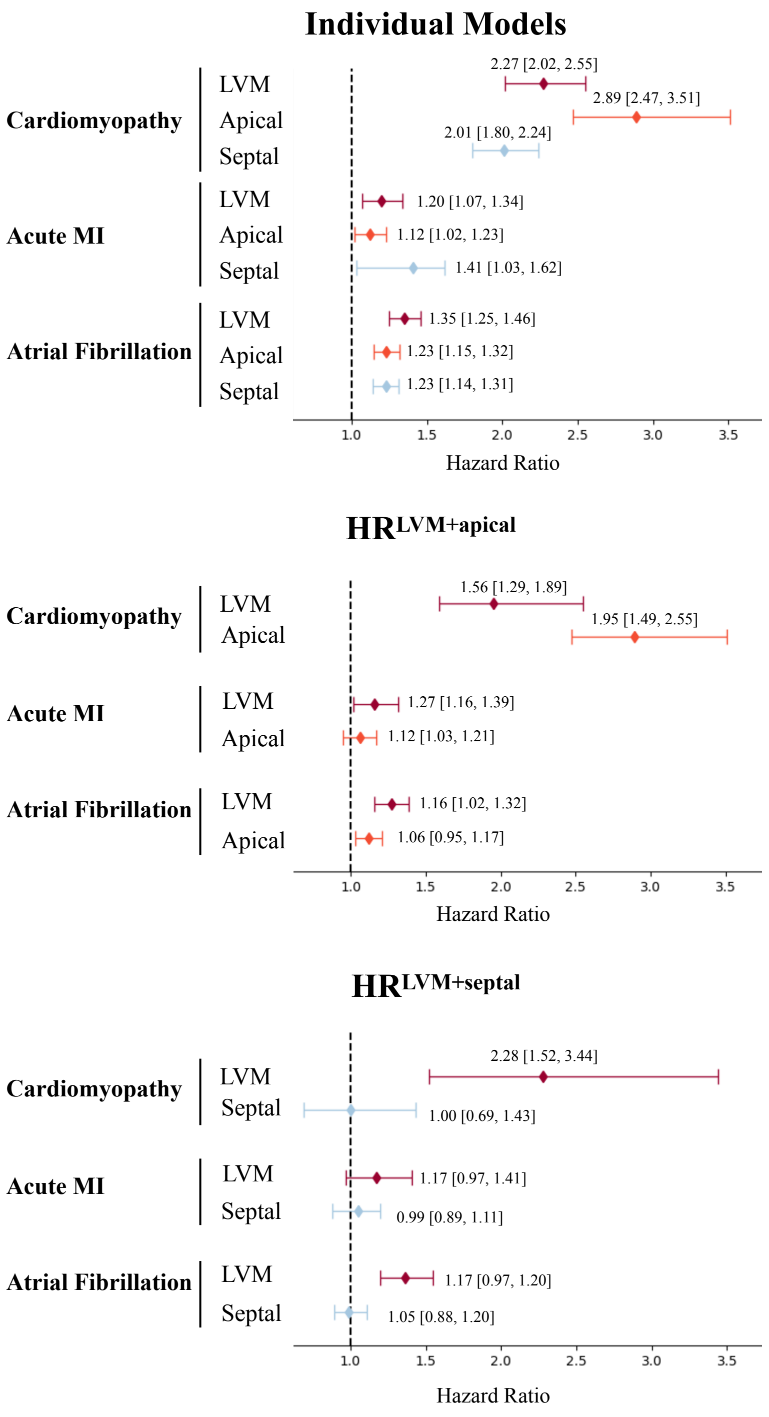
