## Supplemental Figure 1 for "Clinical and genetic associations of asymmetric apical and septal left ventricular hypertrophy"

**Supplementary Information**

**Supplementary Figure 1.** Flow diagram to create the cohort for genetic and phenotypic analysis

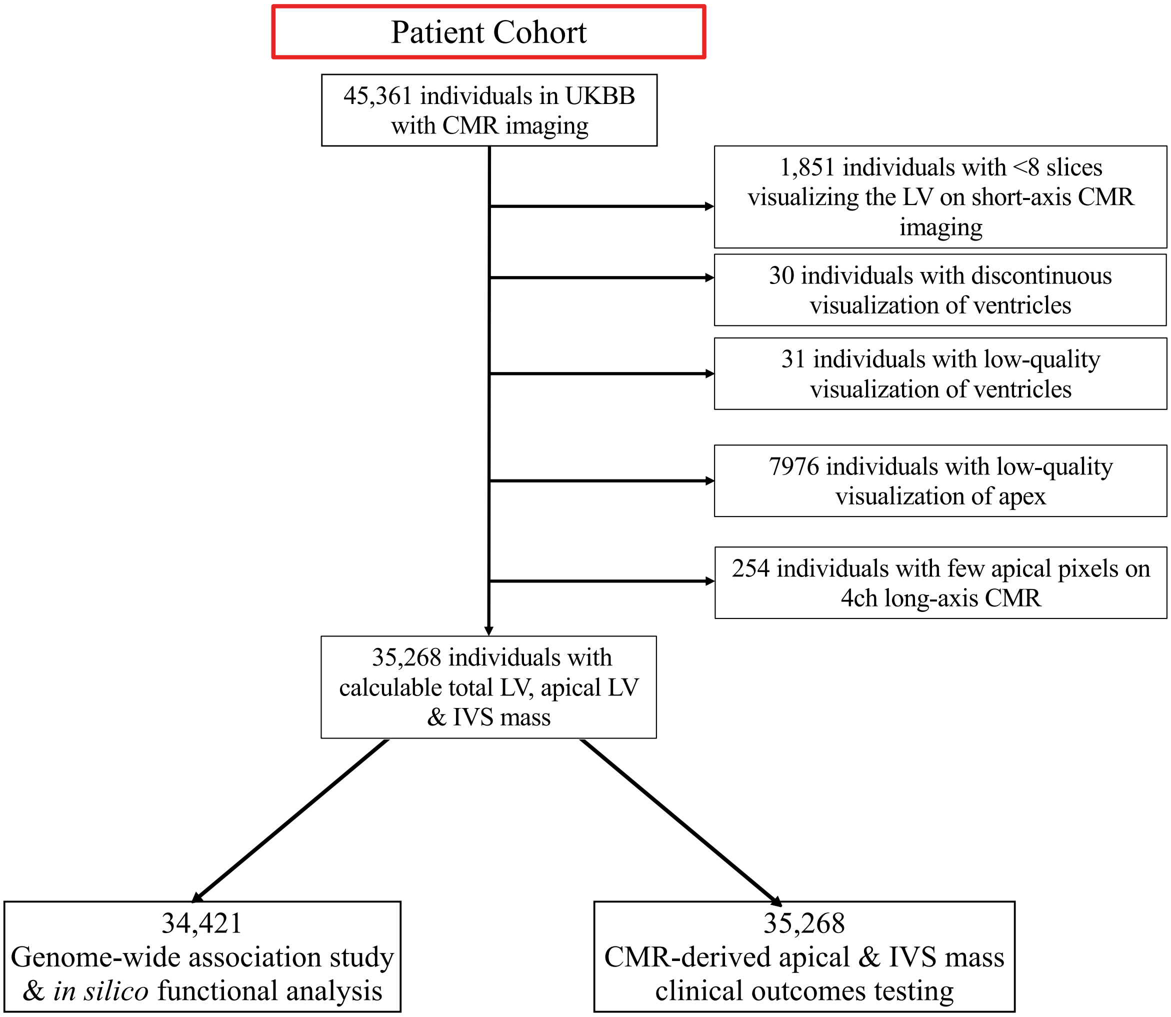
