## Supplemental Table 1 for "Clinical and genetic associations of asymmetric apical and septal left ventricular hypertrophy"

**Supplementary Table 1.** Results of eQTL analysis using FUMA for lead variants in the GWAS of apical and septal mass

| Phenotype | Chr | Candidate Gene | hg37 Position | Ref | Alt | Tissue | p-value |
|---|---|---|---|---|---|---|---|
| Apex | 1 | **CASQ2** | 116,297,758 | A | C | Coronary Artery | 8.51e-10 |
| Apex | 1 | **CASQ2** | 116,297,758 | A | C | Atrial Appendage | 3.95e-8 |
| Apex | 1 | **CASQ2** | 116,297,758 | A | C | Left Ventricle | 1.21e-9 |
| Apex | 6 | **PLN** | 118,876539 | T | C | Aorta | 2.84e-8 |
| Apex | 6 | **PLN** | 118,876539 | T | C | Atrial Appendage | 1.5e-4 |
| Apex | 17 | MAPT-AS1 | 43,948,347 | C | T | Left Ventricle | 7.80e-5 |
| Septal | 3 | **ESYT3** | 138,172,951 | A | G | Coronary Artery | 1.0e-5 |
